# Comparing the Canadian front-of-pack labeling regulations with other mandatory approaches in the Americas and their ability to identify ultra-processed products

**DOI:** 10.1101/2024.11.08.24317012

**Authors:** Nadia Flexner, Fabio S. Gomes, Christine Mulligan, Mavra Ahmed, Laura Vergeer, Jennifer J. Lee, Hayun Jeong, Mary R. L’Abbe

## Abstract

**Background:** Front-of-pack labeling (FOPL) has been implemented in several countries in the Americas, with Chile being the first to introduce a mandatory ‘high in’ warning FOPL in 2016. The Pan American Health Organization (PAHO) food classification criteria, considered a best practice for FOPL regulations, has been adopted by Mexico, Argentina, and Colombia. Canada’s FOPL regulations were recently approved and will take effect in January 2026, but it is unknown how these regulations compare to FOPL regulations that have already been implemented in other parts of the region.

**Objectives:** To compare the Canadian criteria for FOPL regulations with other FOPL criteria implemented in the Americas, and to determine their ability to identify ultra-processed products (UPPs).

**Methods:** Packaged foods and beverages (n=17,094) from the University of Toronto’s Food Label Information and Price (FLIP) 2017 database were analyzed using three FOPL criteria (Canadian, Chilean and PAHO criteria) and the NOVA classification system. The proportions of products that would be subject to displaying a ‘high in/excess’ FOPL and UPPs that would not be subject to FOPL regulations were examined under each system’s criteria. Agreement patterns were modeled using a nested sequence of hierarchical Poisson log-linear models. The Wald statistics for homogeneity were used to test whether proportional distributions differ significantly.

**Results:** Under the Canadian, Chilean and PAHO criteria, 54.4%, 68.4%, and 81.3% of packaged products would be required to display a ‘high in/excess’ FOPL, respectively. Disagreements between the Chilean and the Canadian criteria with PAHO’s were significant, but the greatest disagreement was between the Canadian and PAHO criteria. According to the Canadian, Chilean, and PAHO criteria, 33.4%, 18.4%, 2.3% of UPPs would not be subject to FOPL regulations, respectively.

**Conclusions:** A significant proportion of products that should be subject to FOPL regulations according to the PAHO criteria would not be regulated under Chilean and Canadian criteria, resulting in high proportion of UPPs that would not be subject to FOPL regulations. The Canadian FOPL criteria are the most lenient, with the highest proportion of UPPs that would not display a FOPL. Results can inform improvements for FOPL regulations in Canada, Chile and other countries.

## Introduction

The burden of noncommunicable diseases (NCDs) has steadily increased in recent decades, accounting for the most deaths worldwide and in Canada[1, 2]. Unhealthy diet is a major preventable risk factor for NCDs, responsible for 11 million deaths and 255 million disability-adjusted life-years (DALYs) globally[3, 4]. The availability and overconsumption of ultra-processed food products (UPPs), usually high in nutrients-of-concern (i.e., sodium, sugars, and saturated fats), has increased in recent decades. Furthermore, a growing body of evidence associates high consumption of UPPs with poor diet quality and mental wellbeing, excess body weight, and diet related NCDs[5–9]. Governments have taken different measures, either on a voluntary or mandatory basis, to improve food environments and diets in order to prevent NCDs[10]. These include the implementation of: front-of-pack labeling (FOPL); sodium and sugar reduction targets for key food categories; restrictions on food marketing to children; menu labeling; bans on industrially produced *trans* fatty acids; and sugar-sweetened beverage (SSB) taxes, among others[10].

Nutrient profiling is defined as ‘*the science of classifying or ranking foods according to their nutritional composition for reasons related to preventing disease and promoting health’*[11]. However, the increasing inclusion of food components, such as additives, or the consideration of nutrients to encourage, such as calcium, into nutrient profiling algorithms indicates that it might be more appropriate to refer to them as ‘food classification systems’ (hereinafter also referred to as ‘criteria’)[12]. Adopting a robust, evidence-based food classification system is essential for two key purposes within FOPL regulations. First, to determine which products will be subject to the regulations, and second, to establish the criteria to be adopted to interpret nutrient content data in order to provide clear and easy to interpret nutritional information to consumers[13].

FOPL provides consumers with clear and easy-to-interpret nutritional information in a visual form on the front of the package. Mandatory FOPL, a cost-effective policy to promote healthy diets[14], has been implemented in several countries, mostly in the region of the Americas[15]. In 2016, Chile became the first country in the region to implement a mandatory warning FOPL system, as part of the comprehensive Chilean Nutritional Food Composition and Advertising Law – Law 20.606[16]. This Law also restricts the availability of food and beverage products displaying ‘high in’ warning FOPL in schools, and the advertising and marketing of these products to children (<14 years)[17]. Other countries have followed similar mandatory ‘high in’ or ‘excess’ FOPL approaches, including Argentina[18], Brazil[19], Canada[20], Colombia[21], Israel[22], Mexico[23], Peru[24], Uruguay[25], and Venezuela[26], and many other countries around the world are currently considering its implementation[15, 27–30].

Mandatory ‘high in’ FOPL systems, such as that in Chile, have proven to be effective in prompting consumers to purchase healthier products[31, 32], encouraging food industry reformulation to lower the sugars and sodium content of products[33, 34], and improving diet quality[35]. Initial evaluations of other countries adopting a mandatory ‘high in’ or ‘excess’ FOPL approach show a decrease in the proportion of foods displaying a FOPL following implementation[36], a decrease in purchases of unhealthy food categories[37], and that consumers use and understand the FOPL [22, 25, 38–40]. In contrast, a recent evaluation of the voluntary Health Star Rating (HSR) system, implemented in Australia and New Zealand in 2014, found no effect of the HSR on consumer purchasing behavior[41]. This is consistent with previous evidence from experimental studies[42, 43]. Additionally, low levels of HSR label adoption were observed (25.2%), and HSR-labelled products were more likely to display a 4.0-5.0 rating (healthier alternatives)[41]. This highlights the effectiveness of mandatory versus voluntary approaches and the key differences between nutrient-specific (i.e., ‘high in’ or ‘excess’ FOPL) and summary indicator (i.e., HSR) FOPL systems.

UPPs are defined within the NOVA food classification system[44]. NOVA categorizes foods according to the nature, extent, and purpose of industrial processing. NOVA classifies foods into four primary categories, namely: NOVA 1, unprocessed or minimally processed foods; NOVA 2, processed culinary ingredients; NOVA 3, processed foods; and NOVA 4, UPPs[44]. UPPs, in particular, are recognized for their hyper-palatability, affordability, immediate consumption readiness, long shelf life, and packaging[44]. Food classification systems, such as the one developed by the Pan American Health Organization (PAHO), evaluate foods that fall under NOVA 3 (processed foods) and NOVA 4 (UPPs) due to the association between overconsumption of these products, specifically UPPs, and negative health outcomes[5–9]. The PAHO food classification system is based on robust scientific evidence that informed the World Health Organization (WHO) Population Nutrient Intake Goals to Prevent Obesity and Related NCDs[45]. The PAHO food classification system has been considered a ‘best practice’ for FOPL regulations and has been adopted in Mexico, Argentina, and Colombia[15, 46], and is currently being considered in other countries[15].

Given that the food classification system chosen for FOPL regulation purposes plays a crucial role in determining which products will be subject to regulations, it is important to have a clear understanding of both the scope and distinctions between the chosen criteria and other well-established food classification systems. Additionally, it is important to assess the ability of these food classification systems in identifying and regulating UPPs, as they are significant sources of nutrients-of-concern, and their high consumption has been linked to several NCDs[9].

To the best of our knowledge, no study has previously compared the recently approved Canadian FOPL regulations to similar mandatory approaches adopted or implemented in other countries. Therefore, the objectives of this study were to compare the Canadian criteria for FOPL regulations with other mandatory approaches implemented in the region of the Americas (i.e., the Chilean criteria and the PAHO criteria adopted by Mexico, Argentina, and Colombia), and to determine the ability of each system’s criteria to identify UPPs.

## Methods

### University of Toronto’s Food Label Information and Price (FLIP) database

A descriptive cross-sectional study was conducted using the University of Toronto’s Food Label Information and Price (FLIP) 2017 database. FLIP 2017 is a nationally representative, comprehensive food composition database that includes information on Nutrition Facts table (NFt), universal product codes (UPC), company, brand, product price, list of ingredients, container size, sampling date, and photos of all sides of the package of branded food and beverage products available in the Canadian food supply (n=17,671 unique products). Data collection was carried out in the Greater Toronto Area between July and September 2017. Data were collected from the three leading Canadian grocery retailers (i.e., Loblaws, Sobeys, and Metro), which represented approximately 70% of the Canadian grocery retail market share. The FLIP 2017 database does not include fresh or unpackaged food products. FLIP is comprised of a web-based software and database and a mobile data collector app that facilitates data collection, storing, and analyses of photos of food packages and labels; detailed methods have been previously published[47].

All food products were classified according to Health Canada’s Table of Reference Amounts (TRA) for Food categories[48]. Food products with missing ingredient information and those that would not be subject to FOPL regulations (e.g., alcoholic beverages, meal replacements and nutritional supplements) were excluded. After exclusions, 17,094 food products were included in this analysis.

### Food classification systems for FOPL regulations

Food and beverage products included in this analysis were evaluated under the final FOPL regulations published in *Canada Gazette II*[20], the Chilean thresholds for FOPL regulations (Phase 3)[49], the PAHO food classification system[45] (hereinafter referred to as Canadian, Chilean, and PAHO criteria), and the NOVA food classification system[44]. Details on the criteria used for each of these FOPL regulations are described in **Supplementary Tables S1-3**.

Co-authors of this study have previously published details on steps undertaken for applying the Canadian criteria for FOPL regulations[50]. In brief, the initial step involved assessing food products against established exemption criteria. There are three types of exemption criteria in the FOPL regulations: (a) technical exemptions for food products exempted from displaying a NFt (e.g., raw single-ingredient meats, fresh fruits and vegetables), (b) health-related exemptions for food products that provide health protective effects (e.g., milk, eggs, fruits and vegetables, vegetable oils), and (c) practical exemptions for foods where the ‘high in’ FOPL would be redundant (e.g., honey, butter, salt, syrup). Then, food products that did not meet any of the exemption criteria were assessed for their levels of the targeted nutrients-of-concern (i.e., saturated fat, sodium, and total sugars) (**Supplementary Table S1**). Recently, Health Canada expanded the exemption criteria for FOPL regulations for dairy-related products by lowering the calcium threshold[51]; however, this expansion occurred after these analyses were conducted and are not included in this study.

Under the Chilean criteria, food products were assessed for their content of energy and targeted nutrients-of-concern (i.e., saturated fat, sodium, and total sugars), by 100 g or mL of product. Thresholds have been established for solid and liquid products, separately. Thus, as a first step, we identified solid and liquid food products according to the definition in the Chilean FOPL regulations. In the Chilean criteria all food products that have any added fat, sodium or sugars are subject to FOPL regulations. However, there are some exemptions, which include: foods sold in bulk; infant formulas; infant foods with no added sugars; food supplements; and non-caloric sweeteners (**Supplementary Table S2**).

All food products included in this analysis were categorized under the NOVA food classification system, following the steps previously published by the developers of the NOVA system[44, 52]. NOVA categories were assigned to food products manually by analyzing information from the list of ingredients. This process was initially conducted by a trained researcher (LV), and then a second trained researcher validated the entire dataset (NF). Any discrepancies were discussed with co-authors (FG and MA), and consensus was reached.

Food products categorized as processed (NOVA 3) or UPPs (NOVA 4) were then evaluated under the PAHO criteria. The PAHO criteria evaluate the excessive content of saturated fat, total fat, sodium, and free sugars, as well as the presence of trans fat or sweeteners in food products (**Supplementary Table S3**). Free sugar information is not available in Canadian food labels; therefore, we used previously published free-sugar estimations from our research group, applying the University of Toronto’s Free Sugar Algorithm, a step-by-step methodology[53, 54].

### Statistical analyses

The proportion of products that would be subject to displaying a FOPL was examined under each FOPL system’s criteria. Log-linear models proposed by Tanner & Young[55] were adjusted to examine how the classification of products required to display a FOPL for sodium, sugars and/or saturated fats according to the Canadian and Chilean criteria agree with the ‘best practice’ for the region of the Americas (PAHO criteria[45]). Agreement patterns were modeled using a nested sequence of hierarchical Poisson log-linear models to examine whether the Chilean and the Canadian criteria agree with the PAHO criteria, considering all parameters of each system’s criteria (i.e., saturated fat, sodium, and total sugars for Canada; saturated fat, sodium, total sugars, and energy for Chile; and saturated fat, sodium, free sugars, trans fat, total fat, and other sweeteners for PAHO), and whether they have different levels of agreement/disagreement with the PAHO criteria when considering only nutrients common to all FOPL criteria (i.e., saturated fat, sodium, and sugars).

The Wald statistics for homogeneity were used to test the hypothesis of whether the distributions of products by categories of classification (e.g., exempted; excessive in sodium, sugars, and/or saturated fats; not excessive in any of these three nutrients) according to the different FOPL criteria were equal. The same statistics were used to examine whether the proportional distribution of products, based on the number of nutrients of concern identified as ‘high in/excess’ for sodium, sugars, and/or saturated fats (0 to 3 nutrients) according to each system’s criteria, was equal.

We estimated the proportions of products that were excessive in sodium, sugars, or saturated fats according to the PAHO criteria, but would not display a FOPL for such nutrients under the Canadian and Chilean criteria, with their respective 95% confidence interval. Prevalence ratios and their respective 95% confidence intervals were estimated to verify whether requiring a product to display a FOPL when applying the Canadian and Chilean criteria differed significantly from the PAHO criteria, and to examine the magnitude of such differences if that was the case, considering nutrients common to all criteria and all parameters of each system.

Lastly, we examined whether the proportion of UPPs that would be subject to FOPL regulations (i.e., required to display at least one ‘high in/excess’ FOPL) according to the three different system’s criteria was equal or not. Finally, the odds of a UPP that would be subject to FOPL regulations by each FOPL criteria were estimated and compared considering only nutrients common to all criteria, all nutrient-related parameters of each FOPL criteria, and all parameters of each system’s criteria.

The analyses were conducted in R language and environment for statistical computing version 4.0.1[56].

## Results

### Comparing nutrients common to all FOPL criteria

When restricting the analyses solely to the three nutrient-of-concern (i.e., sodium, sugars, and saturated fats) that are common to the three FOPL system’s criteria, 54.4%, 66.1%, and 77.9% of products would be excessive in at least one nutrient-of-concern under the Canadian, Chilean, and PAHO FOPL criteria, respectively (**Table 1**). Comparing by nutrient-of-concern, the Canadian, Chilean, and PAHO FOPL criteria would identify, respectively, 28.0%, 39.2%, and 48.9% of products as excessive in sodium; 24.3%, 34.0%, and 38.6% as excessive in sugars; and 19.9%, 19.5%, and 32.3% as excessive in saturated fats (**Table 1 and Figure 1**). The proportion of products rated as excessive, not excessive or that are exempted by the criteria for sodium, sugars, or saturated fats, differed significantly between the different systems (p<0.05) (**Figure 1**).

**Table 1.**
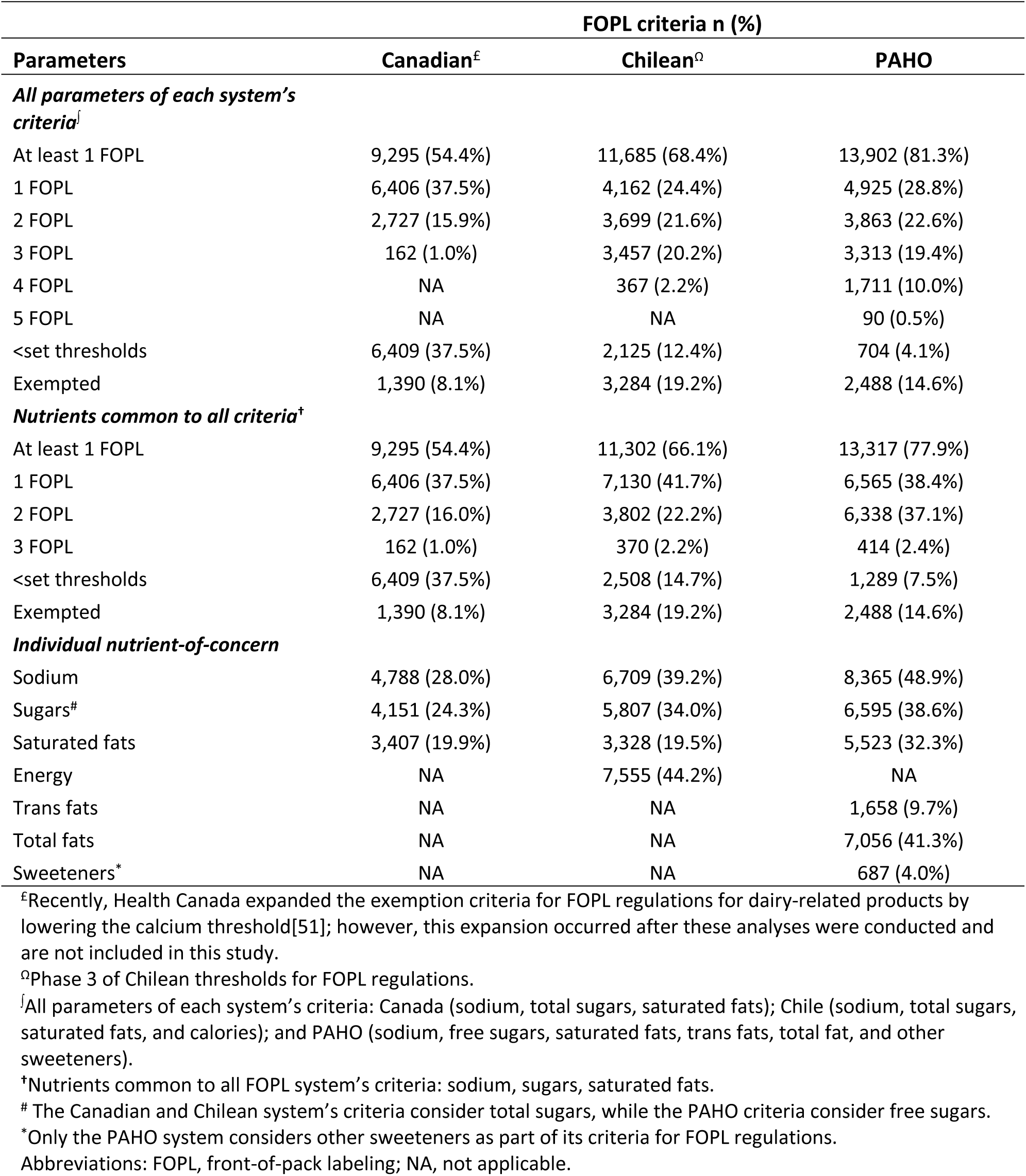
Summary of products that would or would not display a ‘high in/excess’ FOPL according to the Canadian, Chilean and PAHO criteria for FOPL regulations – considering all parameters of each system’s criteria^#^, nutrients common to all criteria**^†^**, and individual nutrient-of-concern (n=17,094)

**Figure 1.**
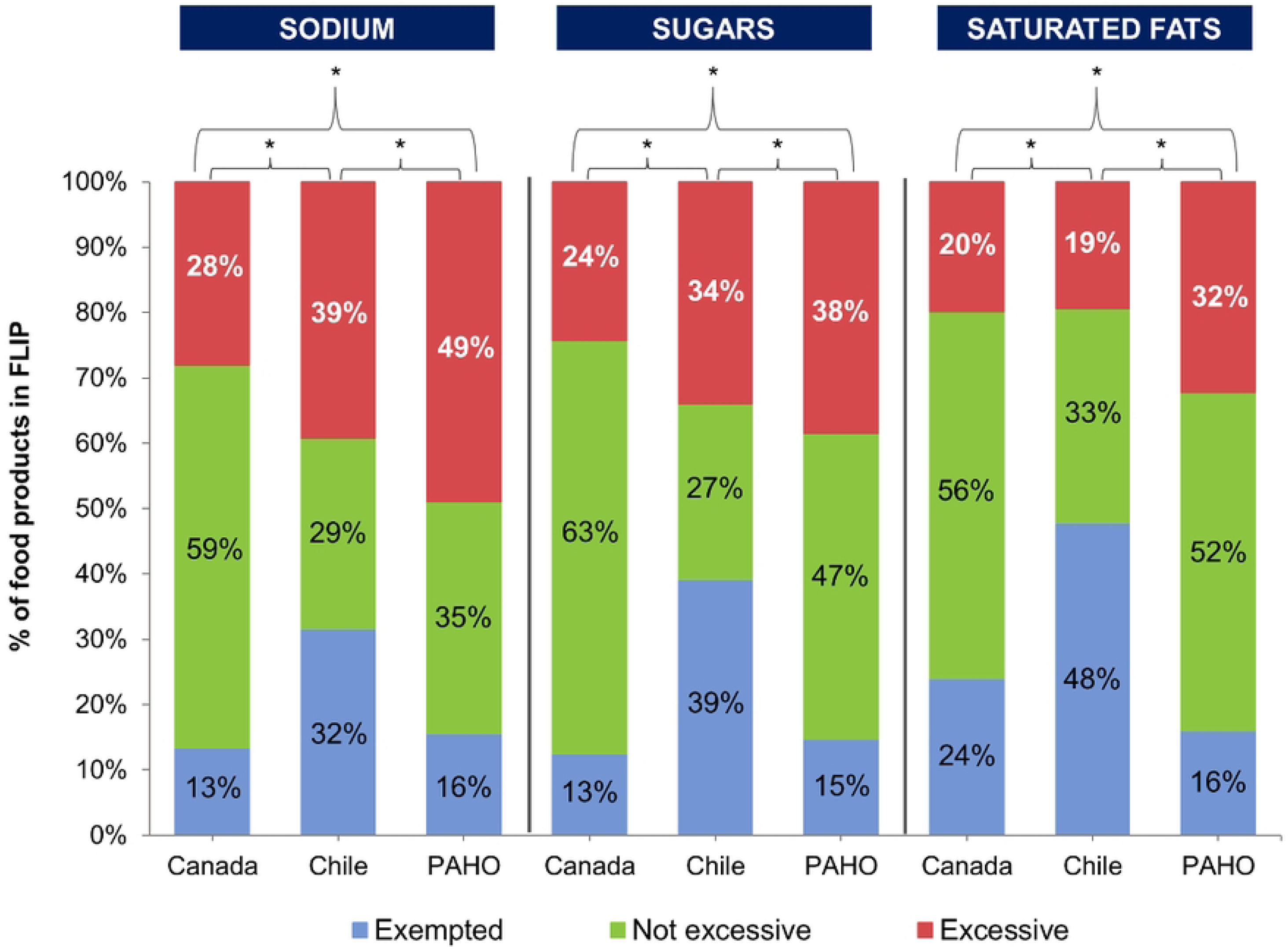
Proportional distribution of products that would or would not display a ‘high in/excess’ FOPL for sodium, sugars^#^, or saturated fats, according to the Canadian, Chilean and PAHO criteria for FOPL regulations (n=17,094) ^#^The Canadian and Chilean system’s criteria consider total sugars, while the PAHO criteria consider free sugars. *Wald statistics for homogeneity indicating when the proportional distributions differ significantly (p<0.05).

The proportions of products that would be subject to 0, 1, 2 or 3 FOPL also differed significantly between the three system’s criteria. The Canadian criteria resulted in the greatest proportion of products that would not display a FOPL for these three nutrients, followed by Chile and PAHO’s criteria (**Figure 2**). **Table 2** shows that 31% and 16% of products that are excessive in sodium, sugars, or saturated fats according to the PAHO criteria would not display a ‘high in/excess’ label under the Canadian and Chilean criteria, respectively. Although both the Canadian and the Chilean criteria result in significant number of products not being subject to FOPL regulations, this is significantly higher under the Canadian criteria than under the Chilean criteria (except for saturated fats, for which the proportion of products not being regulated under the Canadian and Chilean criteria are similar) (**Table 2**).

**Figure 2.**
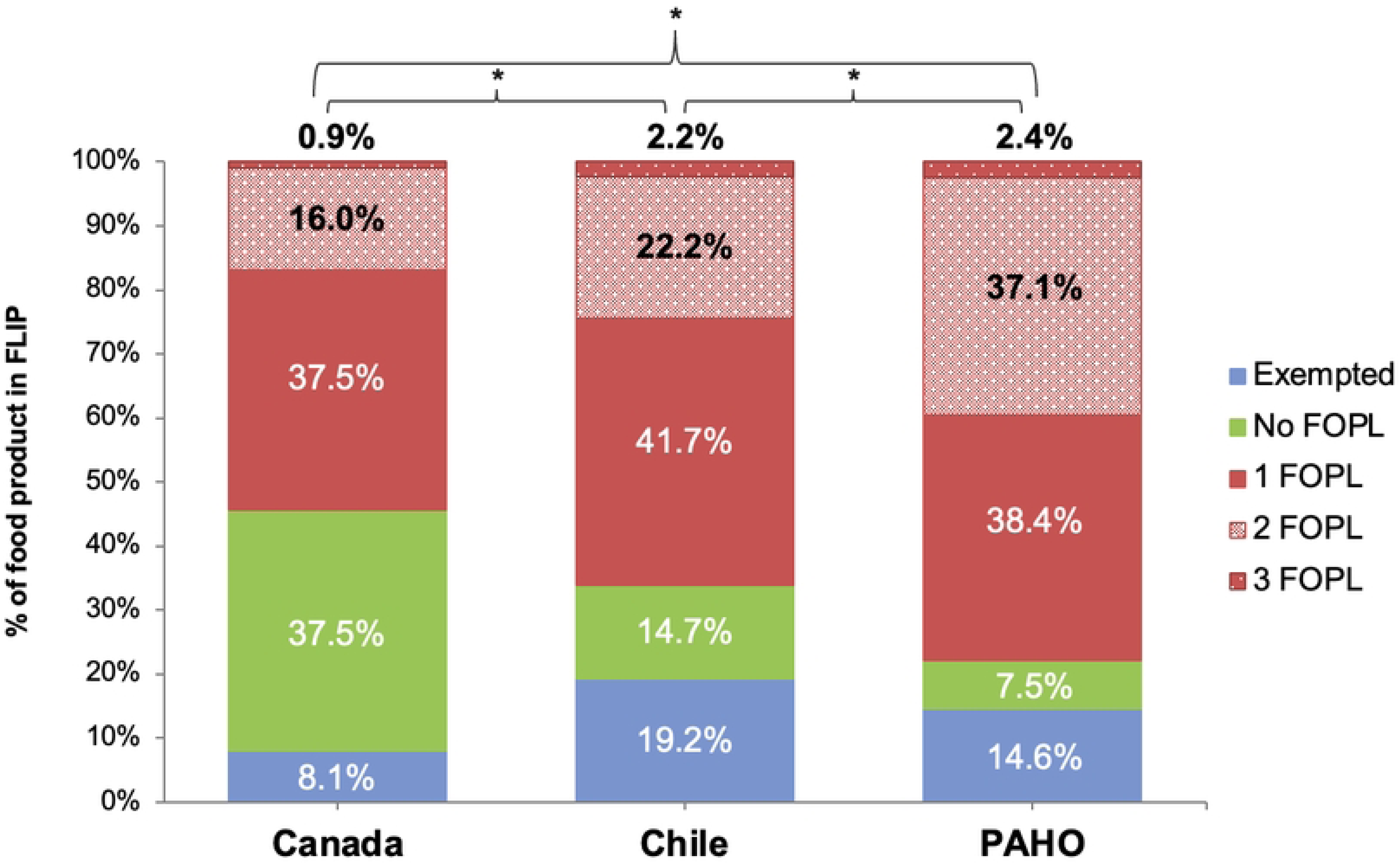
Proportional distribution of products based on the number of nutrients-of-concern identified as ‘high in/excess’ for sodium, sugars^#^, and/or saturated fats according to the Canadian, Chilean and PAHO criteria for FOPL regulations (n=17,094) #The Canadian and Chilean system’s criteria consider total sugars, while the PAHO criteria consider free sugars. *Wald statistics for homogeneity indicating when the proportional distributions differ significantly (p<0.05).

**Table 2.** Prevalence ratios of products that are required to display a ‘high in/excess’ FOPL for sodium, sugars^#^, or saturated fats, or be regulated^Ω^ by the Canadian and the Chilean criteria <u>compared to the PAHO criteria (n=17,094)</u>

|  | PR (95%CI) |  |
| --- | --- | --- |
|  | Canada | Chile |
| Excessive in SODIUM | 0.57 (0.56; 0.58)<sup>†a</sup> | 0.79 (0.78; 0.80)<sup>†b</sup> |
| Excessive in SUGARS<sup>#</sup> | 0.61 (0.60; 0.63)<sup>†a</sup> | 0.88 (0.87; 0.89)<sup>†b</sup> |
| Excessive in SATURATED FATS | 0.62 (0.60; 0.63)<sup>†</sup> | 0.60 (0.59; 0.62)<sup>†</sup> |
| Excessive in SODIUM, SUGARS<sup>#</sup>, OR SATURATED FATS | 0.69 (0.68; 0.70)<sup>†a</sup> | 0.84 (0.83; 0.85)<sup>†b</sup> |
| Regulated<sup>Ω</sup> according to all parameters of each criteria | 0.66 (0.65; 0.67)<sup>†a</sup> | 0.83 (0.82; 0.84)<sup>†b</sup> |

Different superscript letters within a row in the comparison between columns (Canadian vs. Chilean criteria) indicate significant differences (p≤0.05).
<sup>†</sup>Significantly different from 1 (p≤0.05).
<sup>Ω</sup>Required to display at least one ‘high in/excess’ FOPL according to each system’s criteria for FOPL regulations.
<sup>#</sup>The Canadian and Chilean system’s criteria consider total sugars, while the PAHO criteria consider free sugars.
Abbreviations: PR, Prevalence Ratio; 95%CI, 95% Confidence Interval.

Results from the modelling of agreement/disagreement patterns showed that there was an association (null hypothesis of independence rejected by the statistics of deviance) between the Chilean, Canadian, and PAHO FOPL criteria (p<0.001). However, there was no significant agreement in rating products as excessive in sodium, sugars and/or saturated fats, when comparing the Chilean and the Canadian criteria against the PAHO criteria (null hypothesis of agreement was rejected by the statistics of deviance) (p<0.001). In addition, the analyses also demonstrated that the levels of disagreement for the Chilean and the Canadian criteria with the PAHO criteria were not uniform (null hypothesis of homogeneous agreement was rejected by the statistics of deviance) (p<0.001). There was less disagreement between the Chilean and PAHO criteria than between the Canadian and PAHO criteria (null hypothesis of nonhomogeneous agreement was rejected by the statistics of deviance) (p<0.001). This was indicated by the log of coefficients delta_Canada_= 1.73 and delta_Chile_= 2.44 in the nonhomogeneous model (data not shown in **Table 3**), which serves to rank the levels of disagreement and indicate that the lower the coefficient the higher the disagreement. The analyses of the agreement/disagreement patterns using all parameters of each system’s criteria showed similar patterns (**Table 3**).

**Table 3.** Analysis of deviance of nested models for agreement on classification of products based on nutrients common to the Canadian, Chilean and PAHO criteria for FOPL regulations, and on all parameters of each system’s criteria (n=17,094)

| Model | G<sup>2</sup> | DF | p-value<sup>ε</sup> |
| --- | --- | --- | --- |
| **Nutrients common to all criteria<sup>†</sup>** |  |  |  |
| Baseline model $\log(m_{ij}) = u + uPAHO + uCanada + uChile + uCanada*uChile$ (independence) | 40874.8 | 640 | <0.001 |
| Model of agreement of the Canadian and Chilean criteria with the PAHO criteria $\log(m_{ij}) = u + uPAHO + uCanada + uChile + uCanada*uChile + \delta_{ij..l}$ | 28717.7 | 639 | <0.001 |
| Model of homogeneous agreement of the Canadian and Chilean criteria with the PAHO criteria $\log(m_{ij}) = u + uPAHO + uCanada + uChile + uCanada*uChile + \delta_{ij..l}$ | 16397.6 | 638 | <0.001 |
| Model of nonhomogeneous agreement of the Canadian and Chilean criteria with the PAHO criteria $\log(m_{ij}) = u + uPAHO + uCanada + uChile + uCanada*uChile + \delta_{ij..l}$ | 14862.8 | 638 | <0.001 |
| **All parameters of each system's criteria<sup>‡</sup>** |  |  |  |
| Baseline model $\log(m_{ij}) = u + uPAHO + uCanada + uChile + uCanada*uChile$ (independence) | 8455.4 | 3 | <0.001 |
| Model of agreement of the Canadian and Chilean criteria with the PAHO criteria $\log(m_{ij}) = u + uPAHO + uCanada + uChile + uCanada*uChile + \delta_{ij..l}$ | 114.3 | 2 | <0.001 |
| Model of homogeneous agreement of the Canadian and Chilean criteria with the PAHO criteria $\log(m_{ij}) = u + uPAHO + uCanada + uChile + uCanada*uChile + \delta_{ij..l}$ | 4.9 | 1 | <0.05 |
| Model of nonhomogeneous agreement of the Canadian and Chilean criteria with the PAHO criteria $\log(m_{ij}) = u + uPAHO + uCanada + uChile + uCanada*uChile + \delta_{ij..l}$ | 4.9 | 1 | <0.05 |

<sup>ε</sup>Significant evidence to reject the model is correctly specified when p-value is $\leq 0.05$ .
<sup>†</sup>Nutrients common to all FOPL system's criteria: sodium, sugars, saturated fats.
<sup>‡</sup>All parameters of each system's criteria: Canada (sodium, total sugars, saturated fats); Chile (sodium, total sugars, saturated fats, and calories); and PAHO (sodium, free sugars, saturated fats, trans fats, total fat, and other sweeteners).
Abbreviations: DF, Degrees of Freedom; G<sup>2</sup>, Deviance.

Results by Health Canada’s TRA food category are presented in **Supplementary Figure S4.**

### Comparing all parameters of each FOPL system’s criteria

When assessing which products would be regulated (i.e., required to display at least one ‘high in/excess’ FOPL) according to all parameters in the Canadian (sodium, total sugars, saturated fats), Chilean (sodium, total sugars, saturated fats, calories) and PAHO criteria (sodium, free sugars, saturated fats, trans fats, total fats, other sweeteners), 34% and 17% of products required to be regulated by the PAHO criteria would not be regulated by the Canadian and Chilean criteria, respectively (**Table 2**).

### The extent of each FOPL system’s criteria on identifying UPPs

Of the total sample of packaged food products, 72.6% (n=12,414) were classified as UPPs. Using all parameters of the Canadian, Chilean, and PAHO FOPL criteria, 33.4%, 18.4%, and 2.3% of UPPs would not be regulated, respectively. PAHO criteria, even when restricted solely to the three nutrients common to the three systems, left the smallest proportion of UPPs that would not display a FOPL and be restricted by other corresponding regulatory policies. The greatest proportions of UPPs not being regulated were found when the Canadian criteria were applied, followed by the Chilean criteria **(Table 4)**. **Table 4** also shows that the inclusion of ‘other sweeteners’ in addition to the nutrient parameters in the PAHO criteria significantly increased the proportion of UPPs captured by this system’s criteria (p<0.05).

**Table 4.**
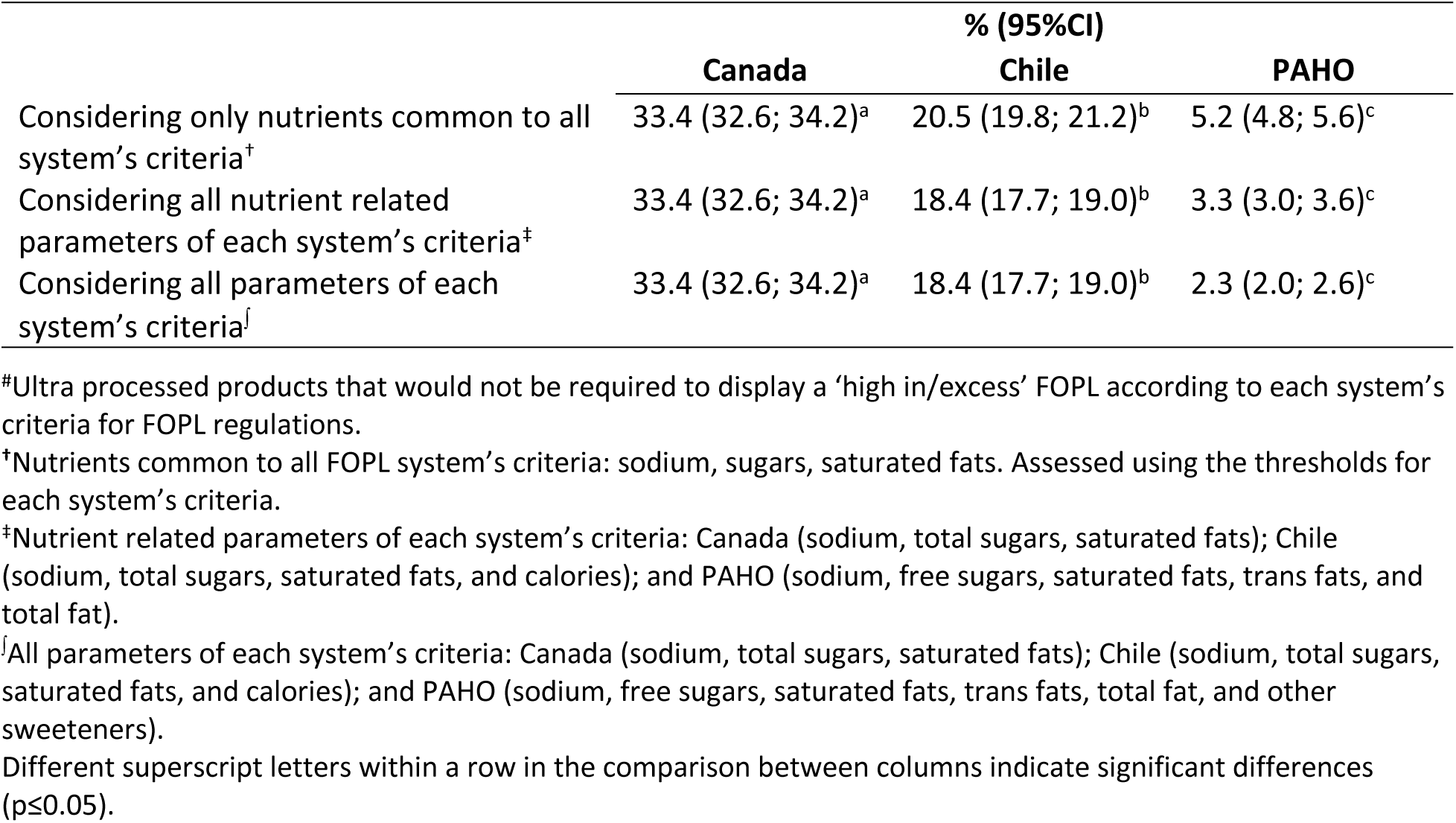
Proportion of ultra processed products that would not be regulated^#^ by the Canadian, Chilean and PAHO criteria for FOPL regulations, and respective 95% confidence intervals (n=17,094)

The results of the binomial logistic regression models showed that the odds of a UPP being regulated under FOPL regulations by the Canadian and Chilean criteria were 21.3 (OR 0.05; 95% CI 0.00, 0.17) and 9.6 (OR 0.10; 95% CI 0.00, 0.23) times lower, respectively, compared to the PAHO criteria. Additionally, the odds of the Canadian criteria regulating UPPs were 2.2 (OR 0.45; 95% CI 0.39, 0.51) times lower than the Chilean criteria (data not shown).

## Discussion

This study aimed to compare the recently published Canadian criteria for FOPL regulations against other criteria implemented in the region of the Americas: the Chilean (Phase 3), the first country to implement a ‘high in’ FOPL system; and the PAHO criteria (adopted by Mexico, Argentina, and Colombia), considered a ‘best practice’ for FOPL regulations. Additionally, we determined the ability of each system’s criteria to regulate UPPs. Our findings indicate that, overall, the Canadian criteria regulate less products than the Chilean and PAHO criteria. The Canadian criteria leave the highest proportion of UPPs unregulated, followed by the Chilean system. When comparing agreement between FOPL criteria, we found more agreement between the Chilean and PAHO criteria than between the Canadian and PAHO criteria.

Furthermore, when looking at individual nutrients-of-concern, our results showed that the Chilean and the Canadian criteria rate fewer products as excessive in sodium, sugars, and saturated fats than the PAHO criteria. Comparisons between the Chilean and Canadian criteria revealed that the Chilean criteria rate more products as excessive in sodium and sugars than the Canadian criteria, with both systems rating similar proportions of products as excessive in saturated fats. This analysis demonstrates that the Canadian thresholds for the nutrients-of-concern are more lenient compared to the Chilean and PAHO criteria. These findings are meaningful given that dietary intakes of nutrients-of-concern among Canadians remain at high levels[57–59], and FOPL regulations have the potential to significantly reduce intakes of targeted nutrients and prevent or delay an important number of diet-related NCD deaths[60]. For instance, *‘bakery products’* and *‘combination dishes’* are the top two contributors of sodium intake in the Canadian diet[61]. Our findings show that an important proportion of these products would not be subject to FOPL regulations under the Canadian criteria, in contrast to the PAHO criteria (**Supplementary Figure S4**). Therefore, to effectively identify products high in nutrients-of-concern, the Canadian criteria for FOPL regulations will need to be informed by a criteria that consider the WHO Population Nutrient Intake Goals to Prevent Obesity and Related NCDs, such as the PAHO criteria, which other countries have adopted, and several others are considering[15, 27–30].

Chile was one of the first countries to establish its own food classification criteria for use in a variety of mandatory policies that promote healthy food choices, as part of its comprehensive Chilean Food Labeling and Advertising Law – Law 20.606[16]. This initiative was in response to the high rates of obesity in Chile, particularly among children, and the high consumption per capita of UPPs[17, 62], a situation similar to that in Canada[62–64]. To date, most post-implementation evidence on mandatory FOPL policies comes from Chile, showing promising initial results[31–35]. Based on the results of this study, which show that the Canadian criteria are less stringent than the Chilean criteria, and considering the comprehensiveness of the Chilean Law in addressing other aspects of the food environment, such as restricting the availability of products with warning labels in schools and limiting food marketing to children (both aspects not implemented in Canada), we can infer that the impact of this policy in Canada may not be of the same magnitude as in Chile. Therefore, monitoring and evaluation of the recently promulgated Canadian FOPL regulations are warranted to measure the impact of the policy.

The PAHO criteria have been adopted and implemented in several countries in the Americas region[46], and is being considered in other countries around the world. For instance, Mexico implemented FOPL regulations in 2020, adopting ‘excess’ octagon warning labels for products exceeding thresholds for calories, sugars, sodium, saturated fats, and trans fats, as well as warning rectangles with the legend ‘avoid/not recommended in children’ for products containing caffeine and non-nutritive sweeteners[65]. The Mexican food classification system is based on the PAHO criteria, with some additional parameters to cover other national public health concerns. These include thresholds for calories and warnings for caffeine given the high consumption of SSBs among Mexicans, including children[65]. Additionally, Mexico added extra thresholds for sodium and did not include thresholds for total fat. Despite these modifications, a recent study assessing the agreement of different food classification criteria applied to packaged products available in the Mexican food supply found almost perfect agreement between the PAHO and Mexican criteria. Additionally, the study identified that 19.9% and 33.4% of packaged foods would be exempted or not required to display a FOPL according to the PAHO and Chilean criteria, respectively. These proportions closely align with our findings (PAHO criteria, 19.9%; Chilean criteria, 31.6%). Our study diverges from the study conducted in Mexico[65] regarding the agreement between PAHO and Chilean criteria, as our findings showed no significant agreement in rating products as excessive in sodium, sugars and/or saturated fats, between these two criteria, most likely due to differences in methodological approaches. Most previous studies comparing food classification criteria[65–67] have used kappa measures which are known to be insensitive to differences between observed and expected patterns of agreement, and they also could not meet necessary assumptions to conduct multiple raters comparisons[55]. We used a nested sequence of hierarchical Poisson log-linear models[55] to avoid these limitations.

The PAHO food classification criteria have been extensively tested to identify food and beverage products excessive in nutrients and ingredients of concern[65, 67–71], and have proven their ability to identify more products excessive in nutrients-of-concern than other FOPL criteria[65, 67–71]. The PAHO system’s novel combination of food processing and nutrient profiling metrics has also been shown to translate outcomes from products to dietary intake levels, facilitating populations’ achievement of WHO healthy diet recommendations[72–75]. It has been recognized as the Americas’ best practice to identify food and beverage products with excessive amounts of nutrients and ingredients of concern[15]. Our results regarding the proportion of products exceeding nutrients-of-concern under the PAHO criteria align with findings from previous studies in other contexts[65, 67–71]. The PAHO system is the only WHO endorsed food classification criteria that also includes ‘other sweeteners’ among the criteria[45], which is a typical marker of UPPs. Our results are consistent with those of Canella et al.[76] on the identification of UPPs using the PAHO criteria. In both studied markets (i.e., Brazil and Canada) approximately 97% of UPPs were identified by the PAHO criteria when using solely the parameters for nutrients in excess, and that proportion rises even closer to 100% when cosmetic additives, including other sweeteners considered by the PAHO criteria, are taken into account[76].

This study has both strengths and limitations that need to be considered when interpreting our findings. We used a nationally representative, branded food composition database to compare Canadian criteria for FOPL regulations against other well-established FOPL criteria. This study used a nested sequence of hierarchical Poisson log-linear approach[55] for testing agreement between the criteria instead of other approaches, such as the kappa index, which are insensitive to differences between observed and expected patterns of agreement/disagreement[55]. Moreover, we analyzed the agreement between food classification criteria by comparing common nutrients and all parameters of each system’s criteria. This comprehensive approach is essential to assess the overall level of disagreement or agreement between criteria, ensuring that various parameters are considered, rather than relying on a few specific parameters. A limitation of our study is that we relied on information reported on the NFt, which allows a permissible margin of ± 20% tolerance when reporting nutrient values[77]. Our analysis assessed mainly packaged food products in the Canadian food supply; thus, proportion of products that would not be subject to FOPL regulations (i.e., fresh products) are underrepresented.

Overall, our findings show that the Canadian and Chilean criteria for FOPL regulations are less stringent and allow significantly more UPPs to omit ‘high in/excess’ labels than the PAHO criteria, with the Canadian criteria showing the most leniency. These findings highlight the importance of implementing robust and independent monitoring mechanisms to assess the effectiveness of the recently published Canadian regulations and suggest that improving the stringency and comprehensiveness of the Canadian FOPL regulations may be warranted to provide the population with the best available policy tools for protecting public health. This study suggests this could be achieved via three modifications to Canada’s FOPL regulations[20]: 1) maintaining the same three nutrients-of-concern used in the current regulations but applying PAHO thresholds for these nutrients, which would reduce to one sixth the proportion of UPPs not subject to FOPL regulations; 2) expanding the criteria to include other nutrients-of-concern, such as total fats, in addition to the existing ones, and using PAHO thresholds, which would reduce to one tenth the proportion of UPPs not subject to FOPL regulations; and finally, 3) full adoption of PAHO criteria, which would reduce to less than one fourteenth the proportion of UPPs not subject to FOPL regulations.

Lastly, supplementary measures may be needed to assist consumers with making healthier food choices and to prompt improvements in the nutritional quality of the Canadian food supply. These measures could include imposing restrictions on the marketing of foods ‘high in’ nutrients-of-concern and the sale of such products in schools, implementing mandatory sodium and sugar reduction targets for key food categories, introducing mandatory calorie labeling on restaurant menus, and aligning FOPL regulations with Canadian Dietary Guidelines recommendations[78].

## Conclusions

The results of this study show that the Canadian criteria for FOPL regulations, followed by that of Chile, are less stringent than the PAHO criteria, and would leave higher proportions of UPPs that would not be subject to FOPL regulations. Neither the Canadian nor the Chilean criteria significantly agree with PAHO’s criteria, but there is greater disagreement between the Canadian criteria and the PAHO criteria than between the Chilean and the PAHO criteria. Our results suggest that Canada’s and Chile’s FOPL regulations could be improved to identify more UPPs and products high in nutrients-of-concern, as has been done by other countries in the Americas. These results can inform future policy decisions in other countries and evaluations of Canadian and Chilean FOPL regulations.

## Data Availability

All relevant data are within the manuscript and its Supporting Information files.

## Authors’ contributions

Conceptualization, NF, FSG, and ML; Data Curation, NF, and FSG; Formal Analysis, NF and FSG; Investigation, NF, CM, MA, LV, JLL, and HJ; Funding Acquisition, ML; Writing – Original Draft Preparation, NF and FSG. All authors critically reviewed and approved the final manuscript. NF is a staff member of the Global Health Advocacy Incubator. FSG is a staff member of the Pan American Health Organization (PAHO). The authors alone are responsible for the views expressed in this article and they do not necessarily represent the views, decisions, or policies of the institutions with which they are affiliated.

## Acknowledgments

The authors would like to thank Alyssa Schermel for her work on managing the Food Label Information and Price (FLIP) database, as well as current and past students who collaborated in different tasks to produce the FLIP database, especially Madyson V. Weippert and Jodi T. Bernstein who worked and published data on free-sugars estimations for the FLIP 2017 database.

## References

1. World Health Organization. Noncommunicable Diseases, Key Facts 2021 [cited 2023 September 13]. Available from: https://www.who.int/news-room/fact-sheets/detail/noncommunicable-diseases.

2. World Health Organization. Noncommunicable diseases country profiles 2018 2018 [cited 2023 September 13]. Available from: https://apps.who.int/iris/handle/10665/274512.

3. Afshin A, Sur PJ, Fay KA, Cornaby L, Ferrara G, Salama JS, et al. Health effects of dietary risks in 195 countries, 1990–2017: a systematic analysis for the Global Burden of Disease Study 2017. The Lancet. 2019;393(10184):1958–72.

4. Stanaway JD, Afshin A, Gakidou E, Lim SS, Abate D, Abate KH, et al. Global, regional, and national comparative risk assessment of 84 behavioural, environmental and occupational, and metabolic risks or clusters of risks for 195 countries and territories, 1990–2017: a systematic analysis for the Global Burden of Disease Study 2017. The lancet. 2018;392(10159):1923–94.

5. Pagliai G, Dinu M, Madarena MP, Bonaccio M, Iacoviello L, Sofi F. Consumption of ultra-processed foods and health status: a systematic review and meta-analysis. Br J Nutr. 2021;125(3):308–18. Epub 2020/08/15. doi: 10.1017/s0007114520002688. PubMed PMID: 32792031; PubMed Central PMCID: PMCPMC7844609.

6. Lane MM, Gamage E, Travica N, Dissanayaka T, Ashtree DN, Gauci S, et al. Ultra-processed food consumption and mental health: A systematic review and meta-analysis of observational studies. Nutrients. 2022;14(13):2568.

7. Askari M, Heshmati J, Shahinfar H, Tripathi N, Daneshzad E. Ultra-processed food and the risk of overweight and obesity: a systematic review and meta-analysis of observational studies. International Journal of Obesity. 2020:1–12.

8. Lane MM, Davis JA, Beattie S, Gómez-Donoso C, Loughman A, O’Neil A, et al. Ultraprocessed food and chronic noncommunicable diseases: A systematic review and meta-analysis of 43 observational studies. Obesity Reviews. 2020.

9. Lane MM, Gamage E, Du S, Ashtree DN, McGuinness AJ, Gauci S, et al. Ultra-processed food exposure and adverse health outcomes: umbrella review of epidemiological meta-analyses. bmj. 2024;384.

10. World Cancer Research Fund International. NOURISHING database: Improve nutritional quality of the whole food supply 2023 [cited 2023 October 20]. Available from: https://policydatabase.wcrf.org/level_one?page=nourishing-level-one#step2=4.

11. World Health Organization Regional Office for Europe. Use of nutrient profile models for nutrition and health policies: meeting report on the use of nutrient profile models in the WHO European Region, September 2021 2022 [cited 2023 October 18]. Available from: https://www.who.int/europe/publications/i/item/WHO-EURO-2022-6201-45966-66383.

12. Martin C, Turcotte M, Cauchon J, Lachance A, Pomerleau S, Provencher V, Labonté M-È. Systematic review of nutrient profile models developed for nutrition-related policies and regulations aimed at noncommunicable disease prevention–An update. Advances in Nutrition. 2023.

13. Pan American Health Organization. Front-of-package labeling as a policy tool for the prevention of noncommunicable diseases in the Americas 2020 [cited 2023 September 3]. Available from: https://iris.paho.org/handle/10665.2/52740.

14. World Health Organization. Tackling NCDs: ’best buys’ and other recommended interventions for the prevention and control of noncommunicable diseases 2017 [cited 2023 October 2]. Available from: https://apps.who.int/iris/handle/10665/259232.

15. Crosbie E, Gomes FS, Olvera J, Rincón-Gallardo Patiño S, Hoeper S, Carriedo A. A policy study on front–of–pack nutrition labeling in the Americas: Emerging developments and outcomes. The Lancet Regional Health - Americas. 2022:100400. doi: 10.1016/j.lana.2022.100400.

16. Ministerio de Salud de Chile. Ley 20.606 - Sobre composicion nutricional de los alimentos y su publicidad 2012 [cited 2023 January 12]. Available from: https://extranet.who.int/ncdccs/Data/CHL_B15_LEY-20606_06-JUL-2012.pdf.

17. Reyes M, Garmendia ML, Olivares S, Aqueveque C, Zacarías I, Corvalán C. Development of the Chilean front-of-package food warning label. BMC public health. 2019;19(1):906.

18. República Argentina. Ley de etiquetado frontal. Promoción de la alimentación saludable Ley 27.642. 2022 [cited 2022 October 12]. Available from: https://www.argentina.gob.ar/justicia/derechofacil/leysimple/salud/ley-de-etiquetado-frontal#:~:text=Est%C3%A1%20prohibida%20la%20publicidad%2C%20promoci%C3%B3n,a%20ni%C3%B1os%2C%20ni%C3%B1as%20y%20adolescentes.

19. Mialon M, Khandpur N, Mais LA, Martins APB. Arguments used by trade associations during the early development of a new front-of-pack nutrition labelling system in Brazil. Public Health Nutrition. 2021;24(4):766–74.

20. Goverment of Canada. Regulations Amending the Food and Drug Regulations (Nutrition Symbols, Other Labelling Provisions, Vitamin D and Hydrogenated Fats or Oils): SOR2022-168 2022 [cited 2022 July 28]. Available from: https://canadagazette.gc.ca/rp-pr/p2/2022/2022-07-20/html/sor-dors168-eng.html.

21. El Congreso de Colombia. Ley No 2120. 2021. [cited 2022 October 5]. Available from: https://www.andi.com.co/Uploads/LEY%202120%20DEL%2030%20DE%20JULIO%20DE%202021.pdf.

22. Shahrabani S. The impact of Israel’s Front-of-Package labeling reform on consumers’ behavior and intentions to change dietary habits. Israel Journal of Health Policy Research. 2021;10(1):1–11.

23. White M, Barquera S. Mexico Adopts Food Warning Labels, Why Now? Health Systems & Reform. 2020;6(1):e1752063.

24. Diez-Canseco F, Cavero V, Alvarez Cano J, Saavedra-Garcia L, Taillie LS, Dillman Carpentier FR, Miranda JJ. Design and approval of the nutritional warnings’ policy in Peru: Milestones, key stakeholders, and policy drivers for its approval. PLOS Global Public Health. 2023;3(6):e0001121. doi: 10.1371/journal.pgph.0001121.

25. Ares G, Antúnez L, Curutchet MR, Galicia L, Moratorio X, Giménez A, Bove I. Immediate effects of the implementation of nutritional warnings in Uruguay: awareness, self-reported use and increased understanding. Public Health Nutrition. 2021;24(2):364–75.

26. FAO, OPS, UNICEF. Etiquetado Nutricional en la Parte Frontal del Envase en América Latina y el Caribe. Nota Orientadora. 2022 [cited 2022 November 4]. Available from: https://iris.paho.org/bitstream/handle/10665.2/56520/9789251367537_spa.pdf?sequence=1&isAllowed=y.

27. Pettigrew S, Coyle D, McKenzie B, Vu D, Lim SC, Berasi K, et al. A review of front-of-pack nutrition labelling in Southeast Asia: Industry interference, lessons learned, and future directions. The Lancet Regional Health-Southeast Asia. 2022.

28. Kroker-Lobos MF, Morales-Juárez A, Pérez W, Kanda T, Gomes FS, Ramírez-Zea M, Siu-Bermúdez C. Efficacy of front-of-pack warning label system versus guideline for daily amount on healthfulness perception, purchase intention and objective understanding of nutrient content of food products in Guatemala: a cross-over cluster randomized controlled experiment. Archives of Public Health. 2023;81(1):108.

29. White-Barrow V, Gomes FS, Eyre S, Ares G, Morris A, Caines D, Finlay D. Effects of front-of-package nutrition labelling systems on understanding and purchase intention in Jamaica: results from a multiarm randomised controlled trial. BMJ open. 2023;13(4):e065620.

30. da Silva Gomes F, Ríos-Castillo I, Correa LRL, Cruzado B, Ares G, Rojas CFU, et al. Effects of front-of-package nutrition labelling systems on objective understanding and purchase intention in Panama: results from a multi-arm parallel-group randomised controlled trial. Public Health Nutrition.1–22.

31. Taillie LS, Bercholz M, Popkin B, Reyes M, Colchero MA, Corvalán C. Changes in food purchases after the Chilean policies on food labelling, marketing, and sales in schools: a before and after study. The Lancet Planetary Health. 2021;5(8):e526–e33.

32. Taillie LS, Reyes M, Colchero MA, Popkin B, Corvalán C. An evaluation of Chile’s Law of Food Labeling and Advertising on sugar-sweetened beverage purchases from 2015 to 2017: A before-and-after study. PLoS medicine. 2020;17(2):e1003015.

33. Reyes M, Smith Taillie L, Popkin B, Kanter R, Vandevijvere S, Corvalán C. Changes in the amount of nutrient of packaged foods and beverages after the initial implementation of the Chilean Law of Food Labelling and Advertising: A nonexperimental prospective study. PLOS Medicine. 2020;17(7):e1003220. doi: 10.1371/journal.pmed.1003220.

34. Quintiliano Scarpelli D, Pinheiro Fernandes AC, Rodriguez Osiac L, Pizarro Quevedo T. Changes in nutrient declaration after the food labeling and advertising law in Chile: a longitudinal approach. Nutrients. 2020;12(8):2371.

35. Fretes G, Corvalán C, Reyes M, Taillie LS, Economos CD, Wilson NLW, Cash SB. Changes in children’s and adolescents’ dietary intake after the implementation of Chile’s law of food labeling, advertising and sales in schools: a longitudinal study. International Journal of Behavioral Nutrition and Physical Activity. 2023;20(1):40. doi: 10.1186/s12966-023-01445-x.

36. Saavedra-Garcia L, Meza-Hernández M, Diez-Canseco F, Taillie LS. Reformulation of Top-Selling Processed and Ultra-Processed Foods and Beverages in the Peruvian Food Supply after Front-of-Package Warning Label Policy. International Journal of Environmental Research and Public Health. 2022;20(1). doi: 10.3390/ijerph20010424.

37. Contreras-Manzano A, White CM, Nieto C, Quevedo KL, Vargas-Meza J, Hammond D, et al. Self-reported decreases in the purchases of selected unhealthy foods resulting from the implementation of warning labels in Mexican youth and adult population. International Journal of Behavioral Nutrition and Physical Activity. 2024;21(1):64.

38. Machín L, Alcaire F, Antúnez L, Giménez A, Curutchet MR, Ares G. Use of nutritional warning labels at the point of purchase: An exploratory study using self-reported measures and eye-tracking. Appetite. 2023:106634.

39. Batis C, Aburto TC, Pedraza LS, Angulo E, Hernández Z, Jáuregui A, et al. Self-reported reactions to the front-of-package warning labelling in Mexico among parents of school-aged children. medRxiv. 2023:2023.07. 26.23293213.

40. Arellano-Gómez LP, Jáuregui A, Nieto C, Contreras-Manzano A, Quevedo KL, White CM, et al. Effects of front-of-package caffeine and sweetener disclaimers in Mexico: Cross-sectional results from the 2020 International Food Policy Study. Public Health Nutrition. 2023:1–30.

41. Bablani L, Mhurchu CN, Neal B, Skeels CL, Staub KE, Blakely T. Effect of voluntary Health Star Rating labels on healthier food purchasing in New Zealand: longitudinal evidence using representative household purchase data. BMJ Nutrition, Prevention & Health. 2022;5(2).

42. Neal B, Crino M, Dunford E, Gao A, Greenland R, Li N, et al. Effects of different types of front-of-pack labelling information on the healthiness of food purchases—a randomised controlled trial. Nutrients. 2017;9(12):1284.

43. Ni Mhurchu C, Volkova E, Jiang Y, Eyles H, Michie J, Neal B, et al. Effects of interpretive nutrition labels on consumer food purchases: the Starlight randomized controlled trial. The American journal of clinical nutrition. 2017;105(3):695–704.

44. Monteiro CA, Cannon G, Levy RB, Moubarac J-C, Louzada ML, Rauber F, et al. Ultra-processed foods: what they are and how to identify them. Public health nutrition. 2019;22(5):936–41.

45. Pan American Health Organization. Pan American Health Organization Nutrient Profile Model 2017 [cited 2023 October 12]. Available from: https://iris.paho.org/handle/10665.2/18621.

46. Global Health Advocacy Incubator. FOPWL regulations around the globe 2023 [cited 2023 October 2]. Available from: https://dfweawn6ylvgz.cloudfront.net/uploads/2023/FOPWL_Regulations.pdf.

47. Ahmed M, Schermel A, Lee J, Weippert M, Franco-Arellano B, L’Abbé M. Development of the Food Label Information Program: A Comprehensive Canadian Branded Food Composition Database. Frontiers in Nutrition. 2022:1319.

48. Health Canada. Table of Reference Amounts for Food 2016 [cited 2021 January 6]. Available from: https://www.canada.ca/en/health-canada/services/technical-documents-labelling-requirements/table-reference-amounts-food.html.

49. Ministerio de Salud de Chile. Decreto 13, Modifica decreto supremo n° 977, de 1996, reglamento sanitario de los alimentos 2015 [cited 2023 October 5]. Available from: https://extranet.who.int/nutrition/gina/sites/default/filesstore/CHL%202015%20Decreto%2013%2C%20Modifica%20decreto%20supremo%20n%C2%BA%20977%2C%20de%201996%2C%20reglamento%20sanitario%20de%20los%20alimentos.pdf.

50. Lee JJ, Ahmed M, Ng A, Mulligan C, Flexner N, L’Abbé MR. Nutrient intakes and top food categories contributing to intakes of energy and nutrients-of-concern consumed by Canadian adults that would require a ‘high-in’ front-of-pack symbol according to Canadian labelling regulations. PLOS ONE. 2023;18(5):e0285095. doi: 10.1371/journal.pone.0285095.

51. Government of Canada. Marketing Authorization to Permit a Lower Calcium Threshold for Exemptions from the Requirement for Prepackaged Products to Carry a Nutrition Symbol in the Case of Cheese, Yogurt, Kefir and Buttermilk: SOR/2024-89 2024 [cited 2024 November 02]. Available from: https://canadagazette.gc.ca/rp-pr/p2/2024/2024-06-05/html/sor-dors89-eng.html.

52. Martinez-Steele E, Khandpur N, Batis C, Bes-Rastrollo M, Bonaccio M, Cediel G, et al. Best practices for applying the Nova food classification system. Nature Food. 2023:1–4.

53. Bernstein JT, Schermel A, Mills CM, L’Abbé MR. Total and free sugar content of Canadian prepackaged foods and beverages. Nutrients. 2016;8(9):582.

54. Weippert MV. Free Sugars in the Canadian Food Supply: Changes from 2013 to 2017 and Modelling the Impact of Sugars Reformulation on the Nutritional Composition of Prepackaged Foods and Beverages: University of Toronto; 2020 [cited 2023 March 8]. Available from: https://hdl.handle.net/1807/121465.

55. Tanner MA, Young MA. Modeling agreement among raters. Journal of the American Statistical Association. 1985;80(389):175–80.

56. R Core Team. The R Project for Statistical Computing 2023 [cited 2023 January 8]. Available from: https://www.r-project.org/.

57. Flexner N, Christoforou AK, Bernstein JT, Ng AP, Yang Y, Fernandes Nilson EA, et al. Estimating Canadian sodium intakes and the health impact of meeting national and WHO recommended sodium intake levels: A macrosimulation modelling study. PLOS ONE. 2023;18(5):e0284733. doi: 10.1371/journal.pone.0284733.

58. Bernstein JT, Christoforou AK, Ng A, Weippert M, Mulligan C, Flexner N, L’Abbe MR. Canadian Free Sugar Intake and Modelling of a Reformulation Scenario. Foods. 2023;12(9):1771.

59. Flexner N, Ahmed M, Mulligan C, Bernstein JT, Christoforou AK, Lee JJ, et al. The estimated dietary and health impact of implementing the recently approved ‘high in’ front-of-package nutrition symbol in Canada: a food substitution scenario modeling study. Frontiers in Nutrition. 2023;10. doi: 10.3389/fnut.2023.1158498.

60. Flexner N, Ng AP, Ahmed M, Khandpur N, Acton RB, Lee JJ, L’Abbe MR. Estimating the dietary and health impact of implementing front-of-pack nutrition labeling in Canada: A macrosimulation modeling study. Frontiers in Nutrition. 2023;10. doi: 10.3389/fnut.2023.1098231.

61. Health Canada. Sodium Intake of Canadians in 2017 2018 [cited 2023 October 12]. Available from: https://www.canada.ca/content/dam/hc-sc/documents/services/publications/food-nutrition/sodium-intake-canadians-2017/2017-sodium-intakes-report-eng.pdf.

62. Pan American health Organization. Ultra-processed food and drink products in Latin America: Trends, impact on obesity, policy implications 2015 [cited 2023 October 5]. Available from: https://iris.paho.org/bitstream/handle/10665.2/7699/9789275118641_eng.pdf?sequence=5&isAllowed=y.

63. Statistics Canada. Health Fact Sheets - Overweight and obese adults, 2018 2019 [cited 2023 September 13]. Available from: https://www150.statcan.gc.ca/n1/pub/82-625-x/2019001/article/00005-eng.htm.

64. Polsky JY, Moubarac J-C, Garriguet D. Consumption of ultra-processed foods in Canada. Health Rep. 2020;31(11):3–15.

65. Contreras-Manzano A, Cruz-Casarrubias C, Munguia A, Jauregui A, Vargas-Meza J, Nieto C, et al. Evaluation of the Mexican warning label nutrient profile on food products marketed in Mexico in 2016 and 2017: A cross-sectional analysis. PLoS Med. 2022;19(4):e1003968. Epub 2022/04/21. doi: 10.1371/journal.pmed.1003968. PubMed PMID: 35442949; PubMed Central PMCID: PMCPMC9067899.

66. Borges CA, Khandpur N, Neri D, Duran AC. Comparing Latin American nutrient profile models using data from packaged foods with child-directed marketing within the Brazilian food supply. Frontiers in Nutrition. 2022;9. doi: 10.3389/fnut.2022.920710.

67. Duran AC, Ricardo CZ, Mais LA, Martins APB. Role of different nutrient profiling models in identifying targeted foods for front-of-package food labelling in Brazil. Public Health Nutrition. 2021;24(6):1514–25.

68. Dickie S, Woods J, Machado P, Lawrence M. Nutrition Classification Schemes for Informing Nutrition Policy in Australia: Nutrient-Based, Food-Based, or Dietary-Based? Current Developments in Nutrition. 2022;6(8):nzac112.

69. Mora-Plazas M, Gómez LF, Miles DR, Parra DC, Taillie L. Nutrition quality of packaged foods in Bogotá, Colombia: A comparison of two nutrient profile models. Nutrients. 2019;11(5):1011.

70. Soares-Wynter S, Aiken-Hemming S-A, Hollingsworth B, Miles DR, Ng SW. Applying nutrient profiling systems to packaged foods and drinks sold in Jamaica. Foods. 2020;9(1):65.

71. Tiscornia MV, Castronuovo L, Guarnieri L, Martins E, Allemandi L. Evaluación de los sistemas de perfiles nutricionales para la definición de una política de etiquetado frontal en Argentina. Revista Argentina de Salud Pública. 2020;12:17-.

72. Machado P, Cediel G, Woods J, Baker P, Dickie S, Gomes FS, et al. Evaluating intake levels of nutrients linked to non-communicable diseases in Australia using the novel combination of food processing and nutrient profiling metrics of the PAHO Nutrient Profile Model. European Journal of Nutrition. 2022:1–12.

73. Köncke F, Toledo C, Berón C, Klaczko I, Carriquiry A, Cediel G, Gomes FS. Estimation of Intake of Critical Nutrients Associated with Noncommunicable Diseases According to the PAHO/WHO Criteria in the Diet of School-Age Children in Montevideo, Uruguay. Nutrients. 2022;14(3):528.

74. Berón C, Toledo C, Köncke F, Klaczko I, Carriquiry A, Cediel G, Gomes FS. Productos procesados y ultraprocesados y su relación con la calidad de la dieta en niños. Revista Panamericana de Salud Pública. 2023;46:e67.

75. Organization PAH. Consumption of Ultra-processed and Processed Foods with Excessive Nutrients Associated with Noncommunicable Chronic Diseases and Unhealthy Diets in the Americas. 2024 [cited 2024 January 8]. Available from: https://iris.paho.org/handle/10665.2/58887.

76. Canella DS, Pereira Montera VdS, Oliveira N, Mais LA, Andrade GC, Martins APB. Food additives and PAHO’s nutrient profile model as contributors’ elements to the identification of ultra-processed food products. Scientific Reports. 2023;13(1):13698.

77. Goverment of Canada. Nutrition labelling compliance test 2024 [cited 2024 August 28]. Available from: https://inspection.canada.ca/en/food-labels/labelling/industry/nutrition-labelling/additional-information/compliance-test.

78. Health Canada. Canada’s Dietary Guidelines for Health Professionals and Policy Makers 2019 [cited 2023 September 18]. Available from: https://food-guide.canada.ca/en/guidelines/.

